# Predicting the impact of low influenza activity in 2020 on population immunity and future influenza season in the United States

**DOI:** 10.1101/2021.08.29.21262803

**Authors:** Kyueun Lee, Hawre Jalal, Jonathan M. Raviotta, Mary G. Krauland, Richard K. Zimmerman, Donald S. Burke, Mark S. Roberts

**Affiliations:** Department of Health Policy and Management, Graduate School of Public Health, University of Pittsburgh, Pittsburgh, Pennsylvania, USA; Public Health Dynamics Laboratory, University of Pittsburgh Graduate School of Public Health; Department of Family Medicine, University of Pittsburgh School of Medicine, Pittsburgh, Pennsylvania, USA; Department of Epidemiology, Graduate School of Public Health, University of Pittsburgh, Pittsburgh, Pennsylvania, USA

## Abstract

**Backgrounds:** The influenza season of 2020-21 was remarkably low, likely due to implementation of public health preventive measures such as social distancing, mask-wearing, and school closure and due to decreased international travel. This leads to a key public health question: what will happen in the 2021-22 influenza season? To answer this, we developed a multi-season influenza model that accounted for residual immunity from prior infection.

**Method:** We built a multi-strain, non-age structured compartmental model that captures immunity over multiple influenza seasons. By the end of the influenza season, we sorted the population based on their experience of natural infection and/or vaccination, which determines the susceptibility to influenza infection in the following season. Because the exact parameters of transmission rates and immunity are unknown, we implemented Bayesian calibration against the observed influenza epidemics (influenza hospitalization rates from 2012 to 2020 in the US) to estimate those parameters. In forward projections, we simulated low influenza activity in 2020-21 season by lowering transmission rate by 20%. Compared to the counterfactual case, in which influenza activity remained at the normal level in 2020-21, we estimated the change in the number of hospitalizations in the following seasons with varying level of vaccine uptake and effectiveness. We measured the change in population immunity over time by varying the number of seasons with low influenza activity.

**Result:** With the low influenza activity in 2020-21, the model estimated 102,000 [95% CI: 57,000-152,000] additional hospitalizations in 2021-22, without change in vaccine uptake and effectiveness. The expected change in hospitalization varied depending on the level of vaccine uptake and effectiveness in the following year. Achieving 50% increase in one of two measures (1.5X vaccine uptake with 1X vaccine efficacy or 1.5X vaccine efficacy with 1X vaccine uptake) was necessary to avert the expected increase in hospitalization in the next influenza season. Otherwise, increases in both measures by 25% averted the expected increase in influenza-hospitalization. If the low influenza activity seasons continue, population immunity would remain low during those seasons, with 48% the population susceptible to influneza infection.

**Conclusion:** We predicted a large compensatory influenza season in 2021-2 due to a light season in 2020-21. However, higher influenza vaccine uptake would reduce this projected increase in influenza.

## Introduction

In response to the public health measures for preventing COVID-19 transmission including social-distancing, school closure, and mask-wearing, and due to reduced international travel, influenza activity was unprecedently low during the influenza season in 2020-21 in the United States (1). According to the CDC surveillance, the cumulative rate of influenza-associated hospitalization by May in 2021 was less than 4 per 100,000 (2). In contrast, during the same period in 2020, the cumulative hospitalization rate had been 70 per 100,000. Total influenza-related deaths decreased by 95% in 2020-21 season, compared to the previous season.

Although low influenza activity resulted in fewer influenza-related hospitalizations and deaths in the 2020-21 season, the impact on the subsequent season is as of now unknown. Those who have never been infected with influenza before (normally infants and children) obtain their first immunity against influenza virus through maternal immunity during pregnancy, vaccination, or natural infection (3-6). The obtained immunity can provide protection against a specific strain of influenza virus beyond the year of infection. During the season with low influenza activity in 2020-21, the population missed the opportunity to establish or boost their immunity for the future influenza season.

Missing opportunities for enhancing immunity during a low influenza season raised the concern of resurgence of influenza in the following season when public health measures are lifted and viral transmission resumes. As COVID-19 containment measures are relaxed around the world, cases of other respiratory viruses whose activity was suppressed under COVID-19 measures in 2020-21 have started to make a resurgence. For example, the epidemics of the respiratory syncytial virus (RSV) in children from Western Australia, which normally peaks in June, started to increase after the county relaxed social distancing measures (7). By the end of December 2020, the number of RSV cases far higher than the seasonal peak in the past. Similarly, Hong Kong had increasing number of outbreaks of acute upper respiratory tract infection after reopening schools (8). The study showed that the total number of school outbreaks involving more than 20 persons from late October to November in 2020 was about seven times the total number of outbreaks from 2017 to 2019. RSV cases started to rise in the U.S. in the summer of 2021, well ahead of the normal pattern (9).

A mathematical model that considers immunity over multiple years is required to estimate the burden of seasonal influenza as a consequence of low influenza activity in 2020-21. Baker RE. and her colleagues used an epidemic model to estimate the impact of COVID-19 nonpharmaceutical interventions on the future dynamics of RSV and influenza in the US (10). The impact of countermeasures such as increasing the coverage and effectiveness of seasonal influenza vaccination on the future epidemic has not been evaluated yet. COVID-19 pandemic and public health measures may have affected the influenza vaccine uptake (11). On the other hand, because vaccine composition is determined based on the surveillance data, low influenza activity can prevent from accurately predicting strains in the following season (12). Hence, this study aimed to estimate the impact of low influenza activity in 2020-21 on the population immunity and the burden of influenza in the following season under various scenarios of vaccine uptake and effectiveness.

## Methods

### Multi-year, multi-strain seasonal influenza model

We built a non-age structured Susceptible-Exposed-Infected-Recovered (SEIR) model which simulated influenza epidemics and consequent population immunity over multiple influenza seasons. We adapted the model structure developed by Hill et al (13) and reproduced the model in R. In brief, the model simulates four main strains of influenza (H1N1, H3N2, B/Yamagata, B/Victoria) starting from 2009. We assumed a perfectly susceptible population in the beginning of 2009-10 season. After 2009-10, the model links epidemiological outcomes in the prior season to the population immunity in the beginning of the “current” season. Exposure history in the past season such as infection with the same strain of influenza, infection with the other B lineage, or vaccination reduced the susceptibility to a specific strain of influenza in a subsequent season. We assumed that vaccine-induced immunity provides protection only for the year of vaccination, whereas infection-induced immunity could remain for at most a single additional influenza season (i.e., 2 seasons total). We provided details of the structure of the seasonal influenza model and implementation of immunity propagation in the model in Appendix 1.

### Model calibration

We conducted Bayesian calibration to estimate the uncertain model parameters related to immunity propagation. Calibration parameters include influenza virus transmissibility by strain (*β*_S_), multipliers for the reduced susceptibility among those who have an exposure history of infection with same strain and the other B lineages *(a, b)*. In addition, to compare the model’s outcome with the observed influenza-related hospitalization rate, we estimated the proportion of total influenza cases that lead to hospitalization (*ε*_*y*_) by season. We described the prior distribution of inferred parameters and the value of fixed parameters in Table 1. As a calibration target, we compared the model’s outcomes to the annual rate of influenza hospitalization from 2012-13 to 2019-2020 in the US (2, 14, 15). We elaborated the details of how we calculated the annual and strain-specific rate of hospitalization from the lab surveillance and total influenza-like illness data in Appendix 2.

**Table 1.**
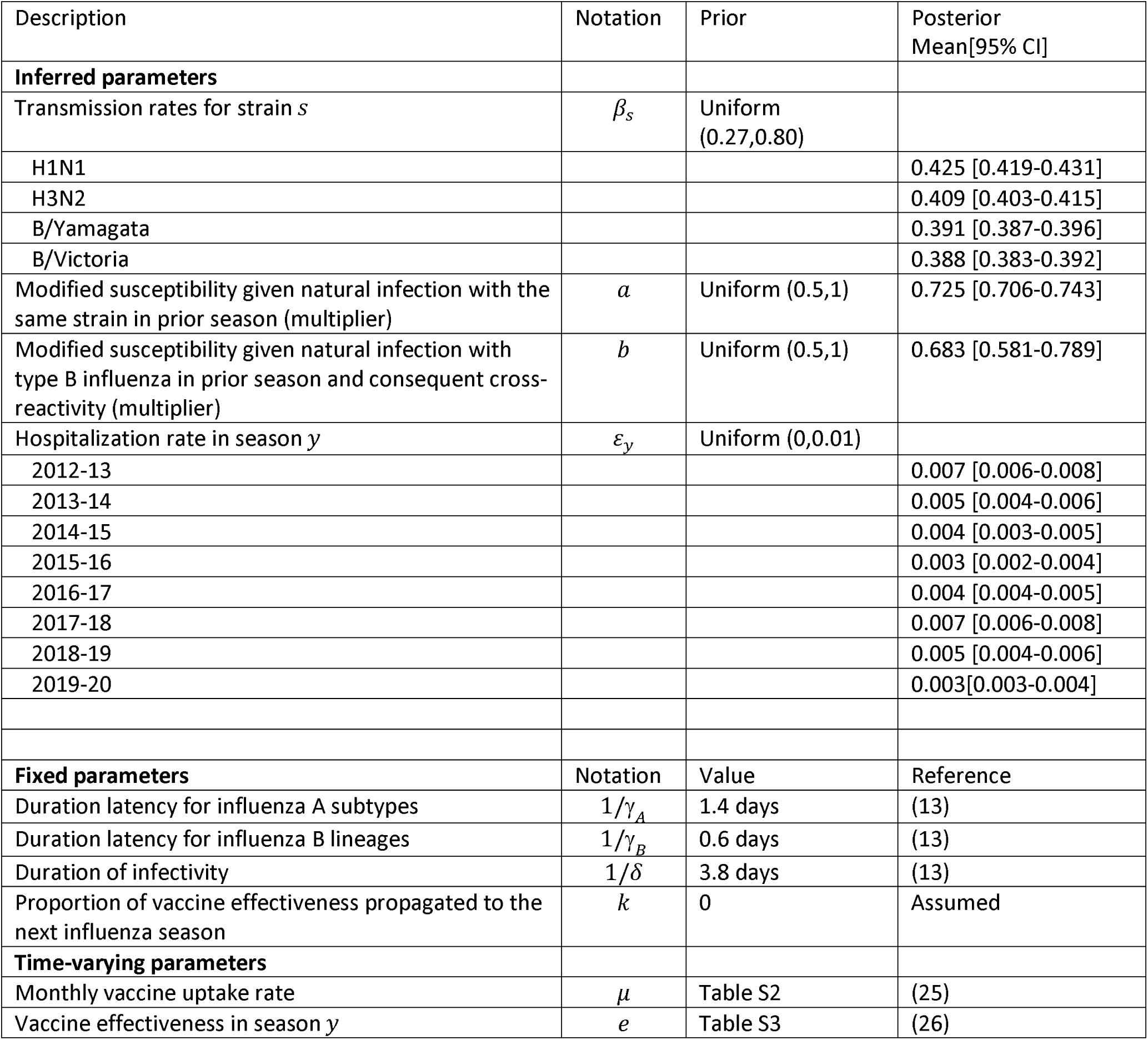
Model parameters

| Description | Notation | Prior | Posterior<br>Mean[95% CI] |
| --- | --- | --- | --- |
| **Inferred parameters** |  |  |  |
| Transmission rates for strain $s$ | $\beta_s$ | Uniform<br>(0.27,0.80) | |
| H1N1 |  |  | 0.425 [0.419-0.431] |
| H3N2 |  |  | 0.409 [0.403-0.415] |
| B/Yamagata |  |  | 0.391 [0.387-0.396] |
| B/Victoria |  |  | 0.388 [0.383-0.392] |
| Modified susceptibility given natural infection with the same strain in prior season (multiplier) | $a$ | Uniform (0.5,1) | 0.725 [0.706-0.743] |
| Modified susceptibility given natural infection with type B influenza in prior season and consequent cross-reactivity (multiplier) | $b$ | Uniform (0.5,1) | 0.683 [0.581-0.789] |
| Hospitalization rate in season $y$ | $\varepsilon_y$ | Uniform (0,0.01) | |
| 2012-13 |  |  | 0.007 [0.006-0.008] |
| 2013-14 |  |  | 0.005 [0.004-0.006] |
| 2014-15 |  |  | 0.004 [0.003-0.005] |
| 2015-16 |  |  | 0.003 [0.002-0.004] |
| 2016-17 |  |  | 0.004 [0.004-0.005] |
| 2017-18 |  |  | 0.007 [0.006-0.008] |
| 2018-19 |  |  | 0.005 [0.004-0.006] |
| 2019-20 |  |  | 0.003[0.003-0.004] |
| **Fixed parameters** |  |  |  |
|  | Notation | Value | Reference |
| Duration latency for influenza A subtypes | $1/\gamma_A$ | 1.4 days | (13) |
| Duration latency for influenza B lineages | $1/\gamma_B$ | 0.6 days | (13) |
| Duration of infectivity | $1/\delta$ | 3.8 days | (13) |
| Proportion of vaccine effectiveness propagated to the next influenza season | $k$ | 0 | Assumed |
| **Time-varying parameters** |  |  |  |
| Monthly vaccine uptake rate | $\mu$ | Table S2 | (25) |
| Vaccine effectiveness in season $y$ | $e$ | Table S3 | (26) |

Because it was computationally expensive to repeatedly running SEIR model over multiple seasons, we instead implemented a meta model in model calibration. A meta model describes the relationship between the simulation model’s inputs (e.g. transmission rates, duration of infectiveness) and outputs (e.g. strain-specific rate of influenza hospitalization) (16). Among many forms of metamodel, artificial neural-network (ANN) meta-model works well when the output of an original model is a non-linear function of its inputs. This method of adopting ANN meta-model for a complex simulation model is called BayCANN (17). We trained an ANN metamodel using independent samples of inputs and corresponding outputs from SEIR and validated the trained ANN metamodel with the test dataset. In Appendix 3, we described the method and result of training and validating ANN in detail. We then implemented the ANN framework within Stan, which is a probabilistic language that performs Hamiltonian Monte-Carlo Markov Chains (18). We evaluated the likelihood of the observed target data given a sampled input parameter to identify distribution of input parameters that can produce model outcomes close to the target. Details of implementing BayCANN with Bayesian calibration using Stan is demonstrated in Appendix 4.

### Examining the change in influenza epidemics in 2021-22 in response to the reduced transmission in 2020/21

We simulated low influenza activity by reducing transmission rate by 10%, 15%, and 20% in 2020-21 and examined the change in influenza epidemic in 2021-22. We also estimated the number of influenza-related hospitalization in the following season under different scenarios of vaccine uptake and effectiveness. In this scenario analysis, we assumed that public health measures to contain COVID-19 transmission reduced the influenza activity by 20% in 2020-21, which produces a season close to the observed activity.

In the status quo scenario (‘historic scenario’ afterwards), the counterfactual case if there were no effect of public health measures on influenza activity in 2020-21, we first projected influenza epidemics in 2020-21 and 2021-22 with the historic influenza transmission rate. In the base case, the influenza transmission rate decreased by 20% in 2020-21 and bounced back to the historic influenza transmission rate (0.425, 0.409, 0.391, 0.388 for H1N1,H3N2, B/Yamagata, B/Victoria, respectively) in 2021-22. In both the historic and base case scenarios, we assumed that the vaccine uptake rate and effectiveness would be the average of corresponding measures in the past influenza seasons from 2009-10 to 2019-20. Hence, by comparing the number of hospitalizations in 2021-22 between two scenarios, we examined the expected increase in influenza-related hospitalizations in 2021-22, after low influenza activity in 2020-21 if there were no change in vaccine uptake rate or effectiveness in 2021-22 season. We further evaluated the sensitivity of the estimated increase in hospitalization with varying level of vaccine uptake and effectiveness in 2021-22 season. We simulated influenza epidemics in 2021-22 with 25%, 50% increased or decreased vaccine uptake and effectiveness from the baseline (average of vaccine uptake and effectiveness in the past influenza season). We first varied one measure at a time while fixing the other variable at the baseline. We then varied both variables simultaneously and measured the change in the number of hospitalizations compared to the historic scenario.

Examining the change in exposure history by the number of low influenza seasons The ongoing COVID-19 pandemic may impose public health and social measures in a long term. We examined how the population’s immunity changes if the low influenza season continues for one, two, or three years since 2020. In the beginning of influenza seasons in 2021, 2022, and 2023, we mapped the entire population into four groups based on their exposure history in the prior year: naïve, natural infection, vaccination, and both. ‘Naïve’ group have no immunity because they did not have infection and vaccination in the prior season, whereas ‘natural infection’, ‘vaccination’, and ‘both’ groups indicate those who have exposure history with infection, vaccination, or both in the prior season.

## Results

### Calibrated seasonal influenza model

Bayesian calibration of seasonal influenza model identified model parameter of which empirical data is insufficient (Table 1). The transmission rates of four influenza strains that were estimated in the historic influenza seasons in the US ranged from 0.391 to 0.425. The estimated transmission rate for the subtype A tends to be higher than the transmission rate of B lineages. Calibration identified 27.5% [95% CI: 25.7-29.4] and 31.7% [95% CI: 21.1-41.9] decreased susceptibility for those who had been infected with the same strain of influenza or the other B lineage. In calibration, we estimated the hospitalization rate among all influenza cases to be 0.3% - 0.7% over the 2012/13-2019/20 seasons. Prior and posterior distribution of parameters are compared in Fig S2. The calibrated seasonal influenza model reproduced the influenza epidemics in the past: the model’s annual rate of influenza-related hospitalization by influenza strain that was close to the corresponding data observed from 2012/13 to 2019/20 (Fig S3). The calibrated model also replicated the dominant influenza strain of each season (Fig S4) in most of historic influenza seasons.

### Change in influenza epidemics in response to the reduced transmission in 2020/21

Lower influenza activity in 2020-21 season leads to higher and earlier seasonal epidemic peak compared to the case when the influenza activity was normal in 2020-21 (Fig 1). With 10% reduced transmission rate, we estimated seasonal peak (defined as when the prevalence of infection is the highest) in 2021-22 to be 0.017 on January 19^th^, 2022. This is 10% higher and 5 days earlier than the expected seasonal peak without the change in influenza activity (0.016). With the 15% and 20% reduced influenza activity in 2020-21, the seasonal peak in 2021-22 increased by 19.7% and 37.7% in 7 and 9 days earlier than expected epidemics with historic influenza pattern.

**Fig 1.**
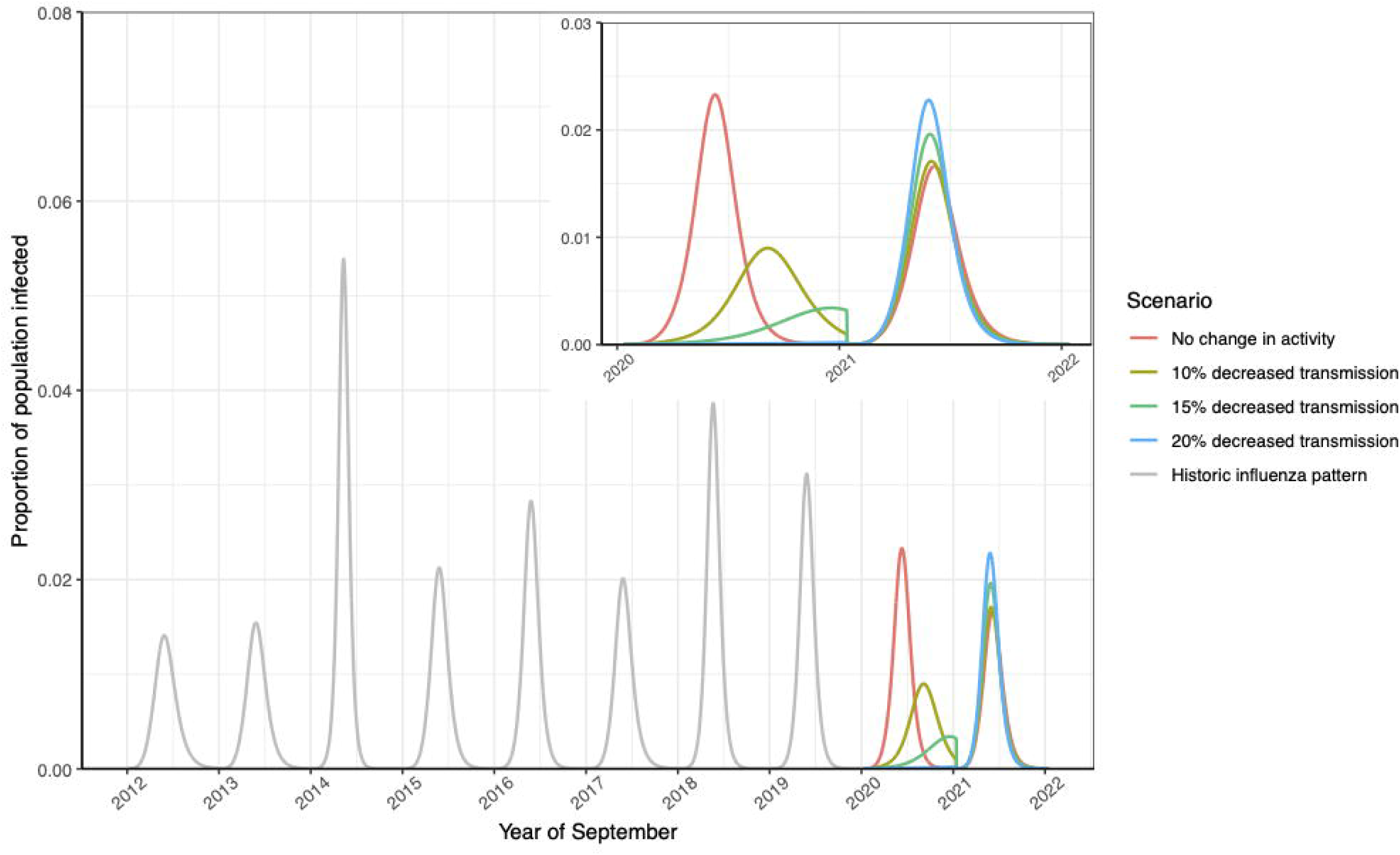
Change in influenza epidemics in response to the reduced transmission in 2020/21. Influenza seasons from 2012 to 2020 were simulated using transmission rates that represents historic influenza patterns (‘Historic influenza pattern’ in grey). The future influenza epidemics from 2020 to 2022 was simulated with the same transmission rates (‘No COVID-19 effect’ in red) or with 10% (olive), 15% (green), and 20% (blue) decreased transmission rates. Trends in 2020-2022 was zoomed in a window.

### Expected increase in the number of influenza-related hospitalizations in 2021/22 season

With low influenza activity in 2020/21, the following influenza season is expected to surge with about 610,000 [95% CI: 500,000-728,000] hospitalizations if vaccine uptake and effectiveness remain stable (Fig 2A). This is 102,000 [95% CI: 57,000-152,000] more hospitalizations compared to the counterfactual case where influenza activity remained same as historic influenza seasons in 2020/21. If the vaccine uptake rate in 2021/22 decreased by 25% and 50% from the average rate in the past influenza seasons, the model estimated the number of influenza-hospitalizations in 2021/22 as high as 709,000 [95%CI: 587,000-842,000] and 808,954 [95% CI: 680,000-950,000] hospitalizations, respectively. The number of hospitalizations in 2021/22 could be lower if the vaccine uptake level reaches higher than the average of past vaccine uptake levels. If vaccine uptake level is 25% and 50% higher than the prior seasons, the number of hospitalizations could decrease to 511,000 [95%CI: 428,000-600,000] and 402,000 [95%CI: 346,000-462,000], respectively. The result was very similar with the varying level of vaccine efficacy in 2021/22 (Fig 2B).

**Fig 2.**
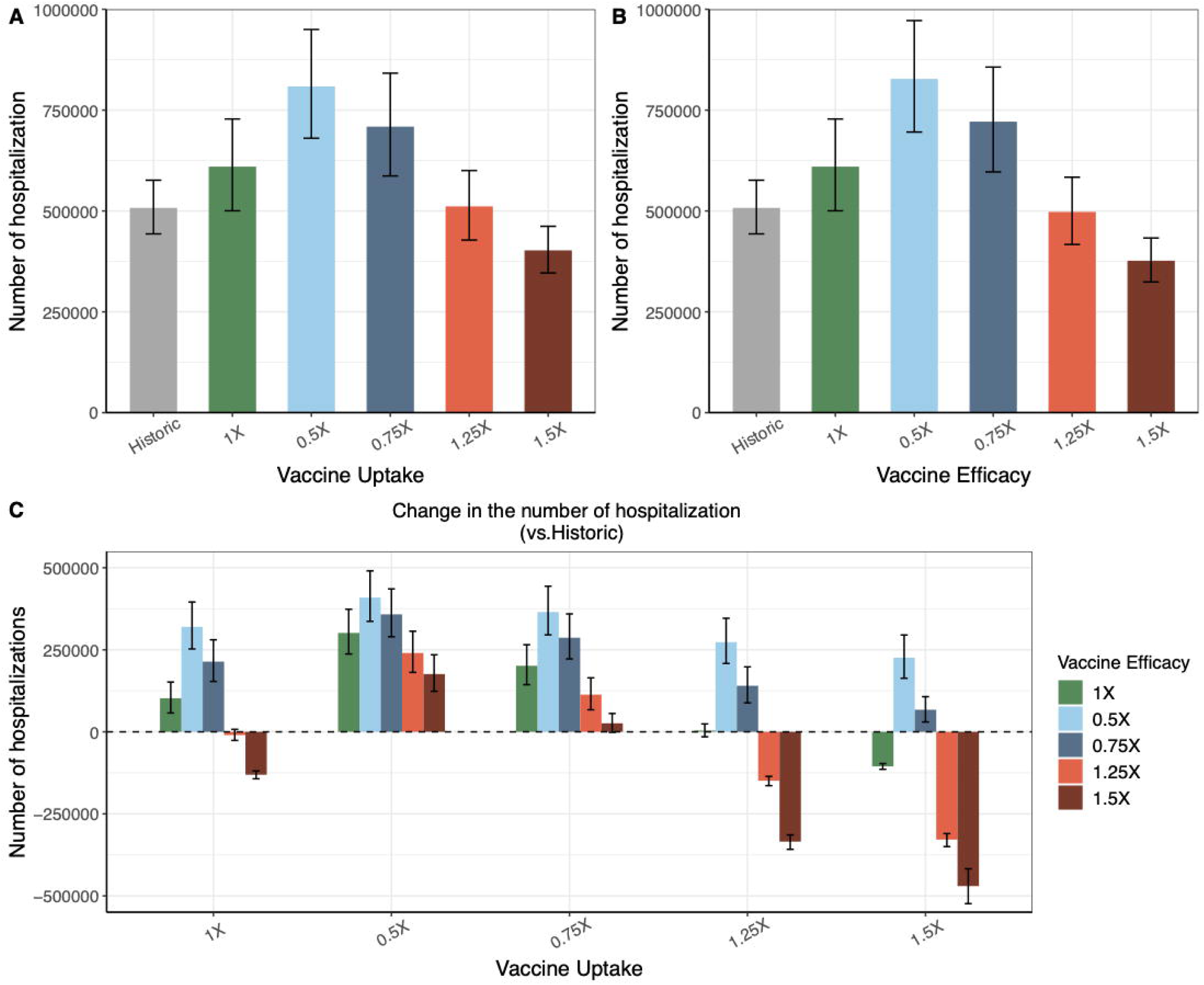
Expected increase in the number of influenza-related hospitalizations in 2021/22 season. ‘Historic’ scenario is the status quo scenario where the influenza activity in 2020/21 remained same as the activity in the historic influenza seasons from 2012/13 to 2019/20. Other scenarios (0.5X-1.5X) assumed low influenza activity in 2020/21 with varying level of vaccine uptake and effectiveness. The label of those scenarios indicates the change in vaccine uptake or effectiveness in 2021/22 from the baseline (defined as the average of measures in the past influenza seasons). (A) The number of influenza-related hospitalization in 2021/22 season with varying level of vaccine uptake in 2021/22 season while the level of vaccine efficacy remained at the baseline. (B) The number of influenza-related hospitalization in 2021/22 season with varying level of vaccine efficacy in 2021/22 season while the level of vaccine uptake remained at the baseline. (C) The expected increase in the number of hospitalizations with varying level of vaccine uptake and efficacy, compared to the number of hospitalizations in ‘historic’ scenario.

In the worst-case scenario, where both vaccine uptake and efficacy were half of the baseline values, our model projected even greater increase in the number of hospitalizations in the next influenza season (409,000 additional hospitalizations, 95% CI: 337,000-490,000) (Fig 2C). Achieving 50% increase in one of two measures (1.5X vaccine uptake with 1X vaccine efficacy or 1.5X vaccine efficacy with 1X vaccine uptake) was necessary to avert the expected increase in hospitalization in the next influenza season. Otherwise, increases in both measures by 25% averted the expected increase in influenza-hospitalization.

### Change in exposure history by the number of influenza seasons

Our model demonstrated that as the influenza seasons with low activity lasts, population immunity for influenza would remain low (Fig 3). If there were no effect of COVID-19 public health measures on influenza activity in 2020/21 (‘No Covid Effect’), 75% of the population have the immunity through infection, vaccination, or both in the beginning of 2021/22 season. The percentage of population immune to at least one strain of influenza remained high afterwards. On the other hand, if influenza activity was low in 2020/21 (‘1-yr effect’), the percentage of the population with immunity decreased to 52% and the rest (48%) had no immunity in the beginning of the following season. The decrease in population immunity was due to decrease in natural infection in the prior season. As the number of years with low influenza activity increases (‘2yr effect’ and ‘3yr effect’), population immunity remained low (52%) until the influenza activity restores back to normal and population obtain immunity through infection.

**Fig 3.**
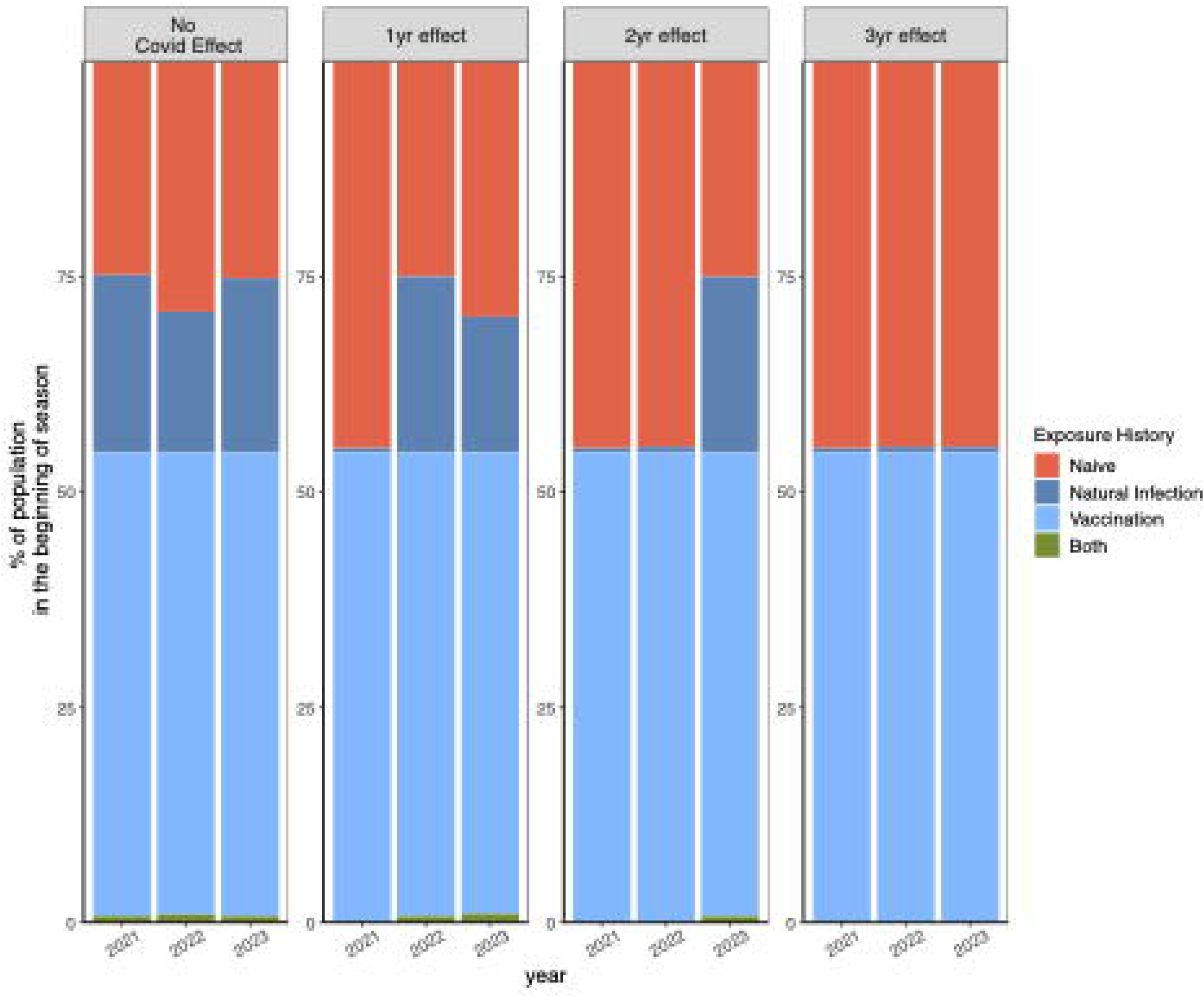
Change in exposure history by the number of influenza seasons with low activity. With no low influenza activity (‘No Covid Effect’), or 1 year, 2-year, or 3-year of low influenza activity since 2020/21 (‘1-yr’, ‘2-yr’, ‘3-yr effect’), population immunity in the beginning of seasons starting from 2021 was mapped to four groups: Naïve, Natural Infection, Vaccination, and Both.

## Discussion

By using a seasonal influenza model that considers immunity propagated over multiple influenza seasons, we showed that the number of influenza-related hospitalizations would increase in the subsequent year when transmission returns to the typical level. The estimated increase, a notable 102,000 hospitalizations, corresponds to 20% increase from the average number of hospitalizations in the past influenza seasons.

We also estimated that as the number of seasons with low influenza activity increased residual immunity in the population decreased. If social distancing measures and school closure continue, and if the influenza activity remains low for multiple seasons, then the susceptible population accumulates over time, which could lead to a large outbreak in a subsequent season. In another modeling study, Baker R.E. and colleagues also found that longer periods of decreased infection are followed by higher and earlier epidemic peaks, due to the increase in susceptibility in the population (10).

Our study highlights the importance of increased influenza vaccination in order to prevent a major outbreak. Achieving a large, 50% increase in either vaccine uptake or vaccine efficacy or 25% increase in both measures was necessary to avert the expected increase in hospitalization in the next influenza season. During the COVID-19 pandemic, the awareness of flu vaccination may have increased and people who have not gotten flu vaccine before may be more willing to receive influenza vaccine this year (11). Preparedness for the potential resurgence of respiratory diseases after COVID-19 measures are relaxed is important. If other infectious diseases whose activity was suppressed under COVID-19 measures resurge in the subsequent year simultaneously, along with ongoing COVID-19 pandemic, this could overwhelm the capacity of the health care system.

This study has several limitations. We only considered immunity from infection to last 2 years. It is possible that immunity built through infection can last longer than two years (19). Uncertainty in the duration of immunity can lead to a large variation in the predicted size of epidemics (20). If infection-induced immunity lasts longer than two years, the predicted size of third and fourth seasons can be smaller than our results. On the other hand, the impact of low influenza activity would be extended beyond the following season, because the reduction in population immunity from infection can alter the susceptibility in farther future influenza seasons. In addition, our model made a simplifying assumption that the effectiveness of vaccine lasts only for one year. This parameter could have been inferred in model calibration. There are two reasons why this parameter was excluded in calibration: first, because the calibration problem becomes unidentifiable with too many uncertain parameters, given the scope of calibration targets and, second, because there was enough evidence on the shorter duration of vaccine-induced immunity compared to infection-induced immunity (21, 22). Lastly, we modeled and estimated overall burden of influenza epidemics without stratifying different risk groups. Upcoming influenza epidemics could disproportionately affect subpopulations, with variation in waning immunity, risk behaviors, and access to healthcare. Current model can be further developed to provide risk group-specific estimate of the expected increase in influenza hospitalizations in the following season.

Strengths of this study include both a multi-year evaluation and modeling multi-strains; such a multi-year, multi-strain model is an advance in the field over single year or single strain models. There are few studies in the current literature that have calibrated influenza model against the observed epidemiological data in the US (23, 24). By combining artificial neural network metamodel and calibration technique, our study inferred uncertain model parameters relevant to viral transmission and immunity propagation that have driven the seasonal influenza epidemics for the past 11 years in the US population. Lastly, our projection on increase in the number of hospitalizations as a consequence of the low influenza activity in 2020 and its variability with vaccine coverage and effectiveness can be valuable information for the preparedness for the upcoming influenza season.

Our study concludes that reduced influenza activity due to public health preventive measures in one season can result in a larger influenza epidemic in the following season if vaccination rates remain stable. Thus, higher influenza vaccine uptake is necessary to reduce the projected increase in burden of influenza in the second season due to reduced residual immunity from lack of seasonal infection in the first season.

## Data Availability

Data availability statement: all the data used for this analysis are publically available. Data from the Center for Disease Control and Prevention for influenza are available at https://www.cdc.gov/flu/index.html.

## Statements

### Ethics approval

As all data used in this study were publicly available, ethics approval was not required.

### Data availability

all the data used for this analysis are publically available. Data from the Center for Disease Control and Prevention for influenza are available at https://www.cdc.gov/flu/index.html.

## Reference

1. Feng L, Zhang T, Wang Q, Xie Y, Peng Z, Zheng J, et al. Impact of COVID-19 outbreaks and interventions on influenza in China and the United States. Nat Commun. 2021;12(1):3249.

2. Influenza Hospitalization Surveillance Network (FluSurv-NET) https://www.cdc.gov/flu/weekly/influenza-hospitalization-surveillance.htm: Centers for Disease Control and Prevention; [

3. Krammer F. The human antibody response to influenza A virus infection and vaccination. Nat Rev Immunol. 2019;19(6):383–97.

4. Kreijtz JH, Fouchier RA, Rimmelzwaan GF. Immune responses to influenza virus infection. Virus Res. 2011;162(1-2):19–30.

5. Nunes MC, Madhi SA. Prevention of influenza-related illness in young infants by maternal vaccination during pregnancy. F1000Res. 2018;7:122.

6. Reuman PD, Ayoub EM, Small PA. Effect of passive maternal antibody on influenza illness in children: a prospective study of influenza A in mother-infant pairs. Pediatr Infect Dis J. 1987;6(4):398–403.

7. Foley DA, Yeoh DK, Minney-Smith CA, Martin AC, Mace AO, Sikazwe CT, et al. The Interseasonal Resurgence of Respiratory Syncytial Virus in Australian Children Following the Reduction of Coronavirus Disease 2019-Related Public Health Measures. Clin Infect Dis. 2021.

8. Fong MW, Leung NHL, Cowling BJ, Wu P. Upper Respiratory Infections in Schools and Childcare Centers Reopening after COVID-19 Dismissals, Hong Kong. Emerg Infect Dis. 2021;27(5):1525–7.

9. Olsen SJ, Winn AK, Budd AP, Prill MM, Steel J, Midgley CM, et al. Changes in Influenza and Other Respiratory Virus Activity During the COVID-19 Pandemic - United States, 2020-2021. MMWR Morb Mortal Wkly Rep. 2021;70(29):1013–9.

10. Baker RE, Park SW, Yang W, Vecchi GA, Metcalf CJE, Grenfell BT. The impact of COVID-19 nonpharmaceutical interventions on the future dynamics of endemic infections. Proc Natl Acad Sci U S A. 2020;117(48):30547–53.

11. Bhatt H. Improving influenza vaccination rates during COVID-19 pandemic - the need of the hour. J Glob Health. 2021;11:03042.

12. Tosh PK, Jacobson RM, Poland GA. Influenza vaccines: from surveillance through production to protection. Mayo Clin Proc. 2010;85(3):257–73.

13. Hill EM, Petrou S, de Lusignan S, Yonova I, Keeling MJ. Seasonal influenza: Modelling approaches to capture immunity propagation. PLoS Comput Biol. 2019;15(10):e1007096.

14. Prevention CfDCa. Disease Burden of Influenza https://www.cdc.gov/flu/about/burden/index.html2009-2020 [

15. Prevention CfDCa. National, Regional, and State Level Outpatient Illness and Viral Surveillance https://gis.cdc.gov/grasp/fluview/fluportaldashboard.html2009-2020 [

16. Jalal H, Dowd B, Sainfort F, Kuntz KM. Linear regression metamodeling as a tool to summarize and present simulation model results. Med Decis Making. 2013;33(7):880–90.

17. Jalal H, Trikalinos TA, Alarid-Escudero F. BayCANN: Streamlining Bayesian Calibration With Artificial Neural Network Metamodeling. Front Physiol. 2021;12:662314.

18. Carpenter B, Gelman, A., Hoffman, M.D., Lee, D., Goodrich, B., Betancourt, M.,. Stan: a probabilistic programming language. Journal of Statististical Software. 2017;76:1–32.

19. Henry C, Palm AE, Krammer F, Wilson PC. From Original Antigenic Sin to the Universal Influenza Virus Vaccine. Trends Immunol. 2018;39(1):70–9.

20. Woolthuis RG, Wallinga J, van Boven M. Variation in loss of immunity shapes influenza epidemics and the impact of vaccination. BMC Infect Dis. 2017;17(1):632.

21. Ferdinands JM, Gaglani M, Martin ET, Monto AS, Middleton D, Silveira F, et al. Waning vaccine effectiveness against influenza-associated hospitalizations among adults, 2015-2016 to 2018-2019, US Hospitalized Adult Influenza Vaccine Effectiveness Network. Clin Infect Dis. 2021.

22. Ferdinands JM, Fry AM, Reynolds S, Petrie J, Flannery B, Jackson ML, et al. Intraseason waning of influenza vaccine protection: Evidence from the US Influenza Vaccine Effectiveness Network, 2011-12 through 2014-15. Clin Infect Dis. 2017;64(5):544–50.

23. Brogan AJ, Talbird SE, Davis AE, Thommes EW, Meier G. Cost-effectiveness of seasonal quadrivalent versus trivalent influenza vaccination in the United States: A dynamic transmission modeling approach. Hum Vaccin Immunother. 2017;13(3):533–42.

24. Sah P, Alfaro-Murillo JA, Fitzpatrick MC, Neuzil KM, Meyers LA, Singer BH, et al. Future epidemiological and economic impacts of universal influenza vaccines. Proc Natl Acad Sci U S A. 2019;116(41):20786–92.

25. FluVaxView, Influenza vaccination coverage. 2009-2020. Centers for Disease Control and Prevention.

26. Seasonal Flu Vaccine Effectiveness Studies 2009-2020. Centers for Disease Control and Prevention.

