## supplement material for "Predicting the impact of low influenza activity in 2020 on population immunity and future influenza season in the United States"

Appendix 1

We built a multi-strain, non-age structured compartmental model that captures immunity over multiple influenza seasons. We adapted and reproduced the model structure from Hill E. et al [ref]. In brief, the model has Susceptible, Exposed, Infected, and Recovered compartments stratified by their vaccination status. Within an influenza season, transitions between compartments occurred at a specified rate. The figure below describes the transitions between compartments:

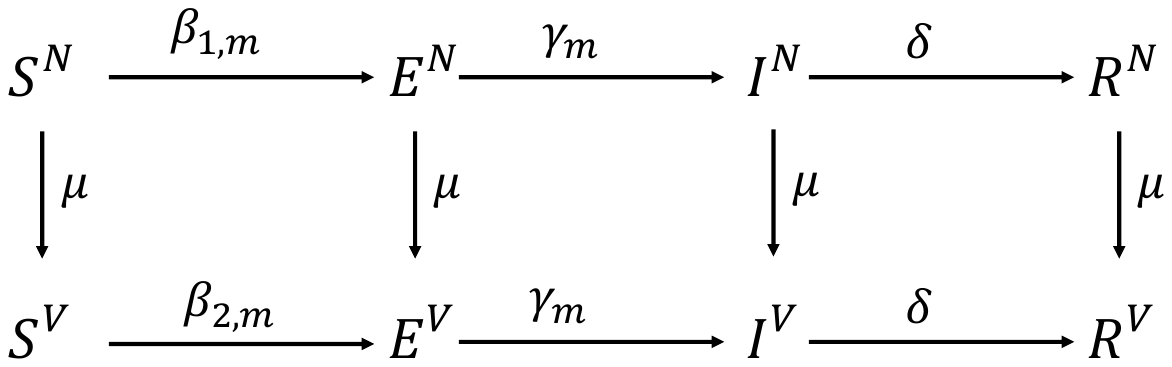

**Figure S1.** Transition between compartments. S: Susceptible, E: Exposed, I: Infected, R:Recovered. Superscript (N, V) in each compartment indicates the vaccinations status. $\beta_{m}$ is a transmission rate for strain m. gamma_m is the rate of developing infectivity. delta is the rate of recovery. Mu is the vaccination rate.

In the beginning of the next season, we mapped the susceptible compartment to ten sub-compartments based on exposure history in the previous season: naïve, exposure history of natural infection (H1N1, H3N2, B/Y, B/V), exposure history of vaccination (Vax), exposure history of both natural infection and vaccination (H1N1-Vax, H3N2-Vax, B/Y-Vax, B/V-Vax). Each exposure history sub-compartment has different modified susceptibility against a strain of influenza virus ($f(h)$). Table below summarizes those modifiers in our model.

|  | H1N1 | H3N2 | B/Y | B/V |
| --- | --- | --- | --- | --- |
| Naïve | 1 | 1 | 1 | 1 |
| H1N1 | 0. 725 | 1 | 1 | 1 |
| H3N2 | 1 | 0. 725 | 1 | 1 |
| B/Y | 1 | 1 | 0. 725 | 0. 683 |
| B/V | 1 | 1 | 0. 683 | 0. 725 |
| Vax | 1 | 1 | 1 | 1 |
| H1N1-Vax | 0. 725 | 1 | 1 | 1 |
| H3N2-Vax | 1 | 0. 725 | 1 | 1 |
| B/Y-Vax | 1 | 1 | 0. 725 | 0. 683 |
| B/V-Vax | 1 | 1 | 0. 683 | 0. 725 |

**Table S1.** Exposure history and corresponding multiplier for the modified susceptibility.

| Month | August | Sep | Oct | Nov | Dec | Jan | Feb | Mar | Apr | May |
| --- | --- | --- | --- | --- | --- | --- | --- | --- | --- | --- |
| 2009/10 | 0.0063 | 0.081 | 0.244 | 0.334 | 0.369 | 0.388 | 0.397 | 0.405 | 0.408 | 0.409 |
| 2010/11 | 0.0063 | 0.081 | 0.244 | 0.344 | 0.373 | 0.401 | 0.415 | 0.422 | 0.429 | 0.43 |
| 2011/12 | 0.011 | 0.078 | 0.245 | 0.336 | 0.366 | 0.388 | 0.405 | 0.413 | 0.417 | 0.419 |
| 2012/13 | 0.022 | 0.092 | 0.246 | 0.341 | 0.380 | 0.411 | 0.430 | 0.440 | 0.446 | 0.449 |
| 2013/14 | 0.023 | 0.096 | 0.276 | 0.373 | 0.406 | 0.437 | 0.454 | 0.458 | 0.464 | 0.468 |
| 2014/15 | 0.026 | 0.098 | 0.288 | 0.389 | 0.428 | 0.454 | 0.465 | 0.476 | 0.480 | 0.483 |
| 2015/16 | 0.023 | 0.098 | 0.273 | 0.366 | 0.400 | 0.428 | 0.445 | 0.454 | 0.458 | 0.463 |
| 2016/17 | 0.024 | 0.100 | 0.275 | 0.369 | 0.403 | 0.432 | 0.449 | 0.458 | 0.462 | 0.467 |
| 2017/18 | 0.024 | 0.093 | 0.246 | 0.324 | 0.355 | 0.383 | 0.4 | 0.4 | 0.414 | 0.417 |
| 2018/19 | 0.024 | 0.101 | 0.286 | 0.38 | 0.417 | 0.449 | 0.469 | 0.481 | 0.488 | 0.492 |
| 2019/20 | 0.027 | 0.113 | 0.313 | 0.413 | 0.454 | 0.486 | 0.506 | 0.513 | 0.517 | 0.518 |

**Table S2.** Cumulative vaccine uptake rate by month from 2009/10 to 2019/20 seasons. We obtained cumulative vaccine coverage in the two age groups 6 months-17 years and 18 years and older from CDC. We calculated average vaccine coverage in the entire population by taking the weighted average of cumulative vaccine coverage using population size in each age group as a weight. Monthly increase in vaccine coverage by month was calculated by taking the difference in cumulative vaccine coverage between two consecutive months. We assumed that monthly vaccine coverage is exponentially distribution with a constant daily vaccination rate.

Reference: CDC FluVaxView, Influenza vaccination coverage. 2009-2020. Available at: <https://www>.cdc.gov/flu/fluvaxview/coverage-by-season.htm

| Season | A(H1N1)pdm09 | A(H3N2) | B/Yam | B/Vic |
| --- | --- | --- | --- | --- |
| 2009/2010 | 0.56 | 0 | 0 | 0 |
| 2010/2011 | 0.45 | 0.52 | 0.6 | 0.6 |
| 2011/2012 | 0.65 | 0.39 | 0.66 | 0.52 |
| 2012/2013 | 0.73 | 0.39 | 0.66 | 0.51 |
| 2013/2014 | 0.54 | 0.54 | 0.52 | 0.52 |
| 2014/2015 | 0.30 | 0.06 | 0.55 | 0.55 |
| 2015/2016 | 0.45 | 0.45 | 0.55 | 0.55 |
| 2016/2017 | 0.33 | 0.33 | 0.52 | 0.56 |
| 2017/2018 | 0.62 | 0.22 | 0.48 | 0.76 |
| 2018/2019 | 0.44 | 0.09 | 0.34 | 0.34 |
| 2019/2020 | 0.3 | 0.3 | 0.45 | 0.45 |

**Table S3.** Vaccine effectiveness from 2009/10 to 2019/20 seasons. If strain-specific effectiveness is available, we used the estimate. If not, we extrapolated missing effectiveness data with only subtype- or lineage-specific data are available. In the year when subtype- or lineage-specific data are not available, we used overall vaccine effectiveness.

Reference: CDC Seasonal Flu Vaccine Effectiveness Studies 2009-2020. Available at: https://www.cdc.gov/flu/vaccines-work/effectiveness-studies.htm

Appendix 2

We estimated the influenza hospitalization rate attributed to each strain of influenza by applying strain composition from the lab surveillance data to the total number of influenza hospitalizations and divide it by the population size in the US.

|  | Strain distribution | | | | |  | Number of influenza hospitalizations by strain | | | |
| --- | --- | --- | --- | --- | --- | --- | --- | --- | --- | --- |
| SEASON | H1N1 | H3N2 | B/V | B/Y | Hospitalizations | | H1N1 | H3N2 | B/V | B/Y |
| 2009-2010 | 0.99 | 0.00 | 0.01 | 0.00 | 265000 | | 262861.0 | 574.5 | 1376.8 | 187.7 |
| 2010-2011 | 0.25 | 0.41 | 0.31 | 0.02 | 290000 | | 73648.8 | 119878.2 | 90741.9 | 5731.1 |
| 2011-2012 | 0.20 | 0.57 | 0.12 | 0.11 | 140000 | | 28311.3 | 80297.8 | 16323.3 | 15067.7 |
| 2012-2013 | 0.03 | 0.58 | 0.14 | 0.25 | 570000 | | 17825.3 | 330085.4 | 79952.1 | 142137.2 |
| 2013-2014 | 0.70 | 0.10 | 0.07 | 0.13 | 350000 | | 246404.7 | 33259.7 | 25320.8 | 45014.8 |
| 2014-2015 | 0.00 | 0.71 | 0.10 | 0.18 | 590000 | | 2183.5 | 417445.8 | 61333.4 | 109037.2 |
| 2015-2016 | 0.61 | 0.18 | 0.07 | 0.15 | 280000 | | 169538.2 | 49287.0 | 19260.0 | 41914.9 |
| 2016-2017 | 0.02 | 0.78 | 0.06 | 0.14 | 500000 | | 12369.4 | 388066.4 | 28013.8 | 71550.3 |
| 2017-2018 | 0.11 | 0.64 | 0.03 | 0.22 | 810000 | | 93068.0 | 518055.9 | 21312.1 | 177564.1 |
| 2018-2019 | 0.54 | 0.43 | 0.02 | 0.01 | 490000 | | 263711.7 | 209513.5 | 11301.3 | 5473.5 |
| 2019-2020 | 0.58 | 0.05 | 0.36 | 0.01 | 400000 | | 230566.0 | 21310.7 | 145782.1 | 2341.3 |

**Table S4.** Strain distribution and total number of influenza-related hospitalizations.

Reference:

CDC Estimated burden of seasonal influenza. Available at: <https://www.cdc.gov/flu/about/burden/past-seasons.html>

CDC FluView Seasonal influenza surveillance data. Available at: <https://gis.cdc.gov/grasp/fluview/fluportaldashboard.html>

Appendix 3.

Training and validating ANN metamodel

We first prepared training and validation dataset by randomly sampling parameter sets and corresponding outcomes from SEIR model. We took the logarithm of the outcomes to have a distirubtion of outcomes close to a normal distribution. We standardized the inputs and outputs so that the values range from -1 to 1. We then fitted an artificial neuro-network model in the training dataset using Keras library in R (https://keras.rstudio.com). The constructed model had six hidden layers and 200 nodes in each hidden layer. Because the inputs and outputs ranged from -1 and 1, we used hyperbolic tangent function as an activation function. We minimized the mean squared errors using a stochastic gradient descent algorithm (Adam optimizer). Adam optimization is a stochastic gradient descent method that is based on adaptive estimation of first-order and second-order moments. Below compares the raw and predicted outcomes from SEIR model and ANN metamodel, respectively in the training dataset and validation dataset.

Appendix 4.

We assumed that prior distribution of model parameters is uniform distribution. The range of prior distribution for influenza transmission rates was set based on the range of rates used in other studies. We varied the modifier for the susceptibility given exposure history from 0.5 to 1. We initially varied from 0 to 1 and found that it renders parameters unidentified in calibration. We made a conservative assumption on the propagated immunity by taking 0.5-1 range instead of 0-0.5 as a modifier.

Within stan, we implemented the trained artificial neural network metamodel for sampled input parameters. We back-transformed the predicted outcomes to compare them with the observed data (calibration target). Stan requires assumptions on the distribution on the observed target data in evaluating their likelihood given the sampled parameter. We assumed that the target data (annual rate of influenza-related hosptializations) are Poisson-distributed. By the nature of Poisson distribution, this assumption will weigh more on the small rate than the large rate because the mean and the variance are equal in Poisson distribution. We added a constraint to the calibration: all parameters in the posterior distribution should simulate seasonal peak before March 1^st^. In order to add this constraint to the calibration and treat all parameters that simulate the peak before March 1^st^ equally, we selected a random peak date as a target and transformed simulated peak date to the target date if parameter meets the condition (peak before March 1^st^). Setting a normal distribution around the target date with a small standard error allows the calibration process to strictly select parameters that generate the seasonal peak before March 1^st^. We set the number of interations as 100,000 and had 4 chains to see whether the outcome converges with different initial values.

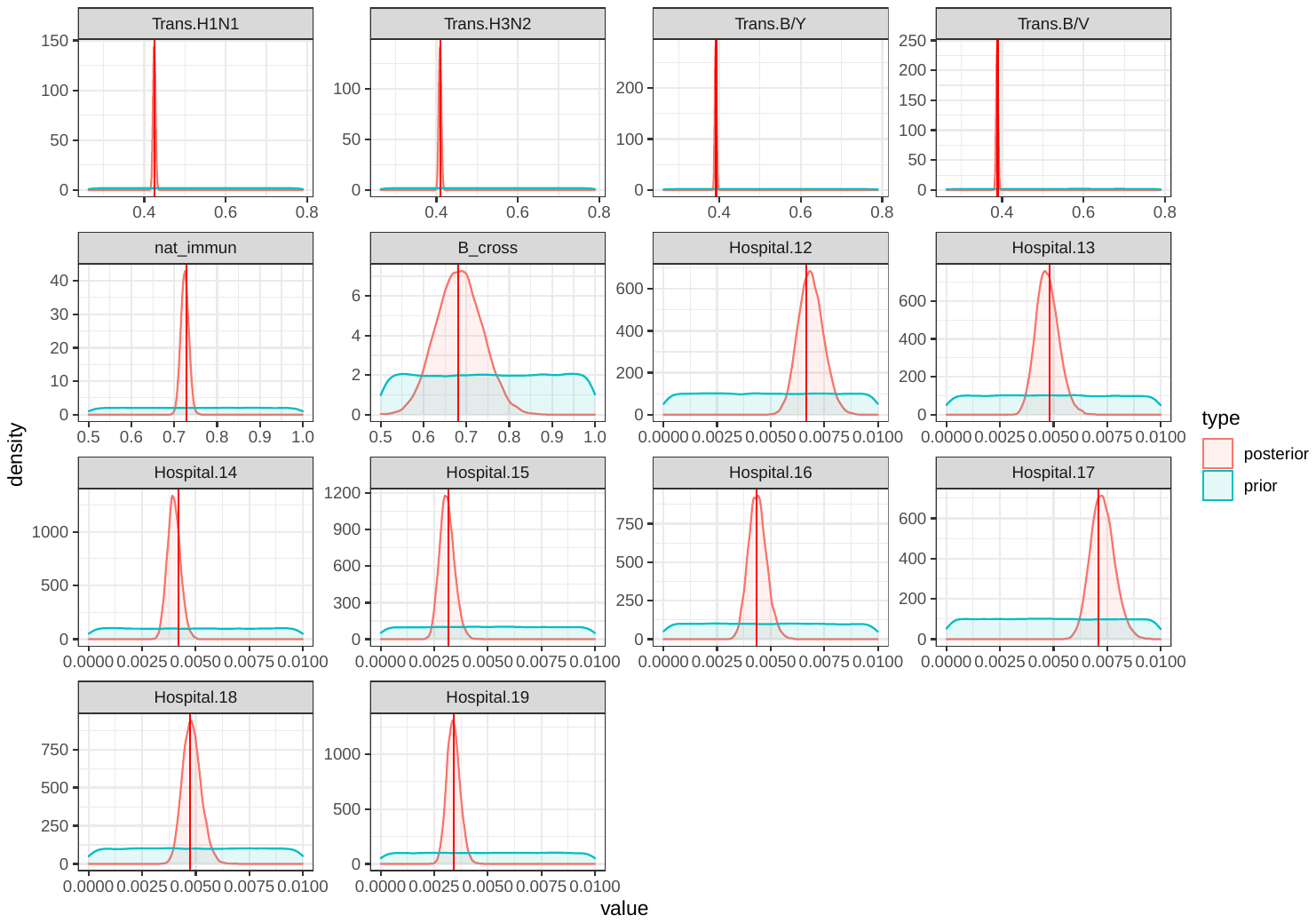

**Fig S2. Posterior distribution of seasonal influenza model parameters.** The distribution in blue indicates prior distribution before model was calibrated to the empirical data on influenza epidemics from 2012/13-2019/20 in the US. The distribution in red indicates posterior distribution of model parameters that was updated from the prior distribution through Bayesian calibration. The red vertical line shows the estimated maximum a posteriori of the posterior distribution.

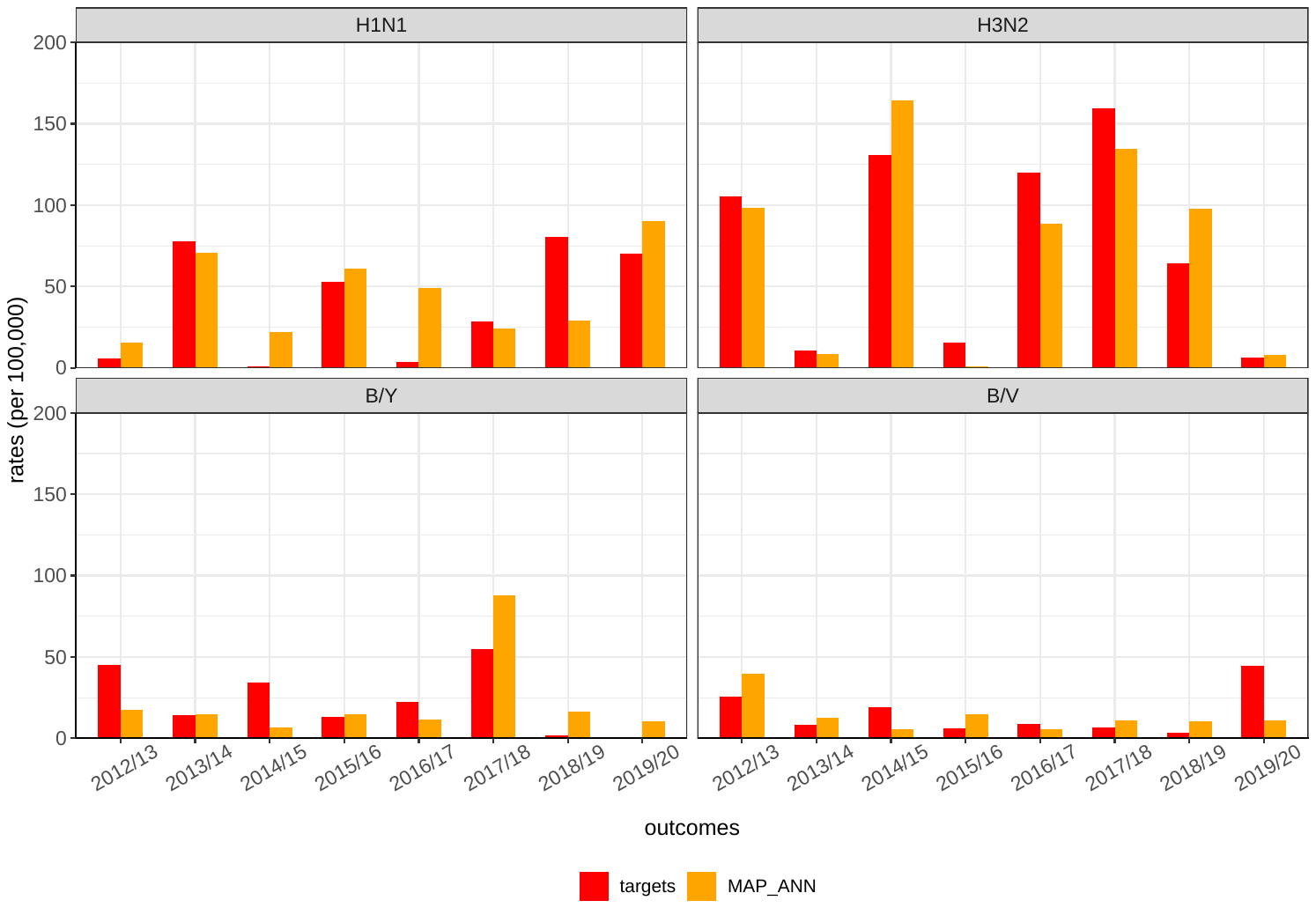

**Fig S2. Comparison of simulated and observed annual hospitalization rate by influenza strain from 2012/13 to 2019/20 season.** Red bar indicates the observed annual rate of influenza hospitalization by influenza strain. Orange bar indicates the corresponding model outcomes simulated by using the maximum a posteriori.

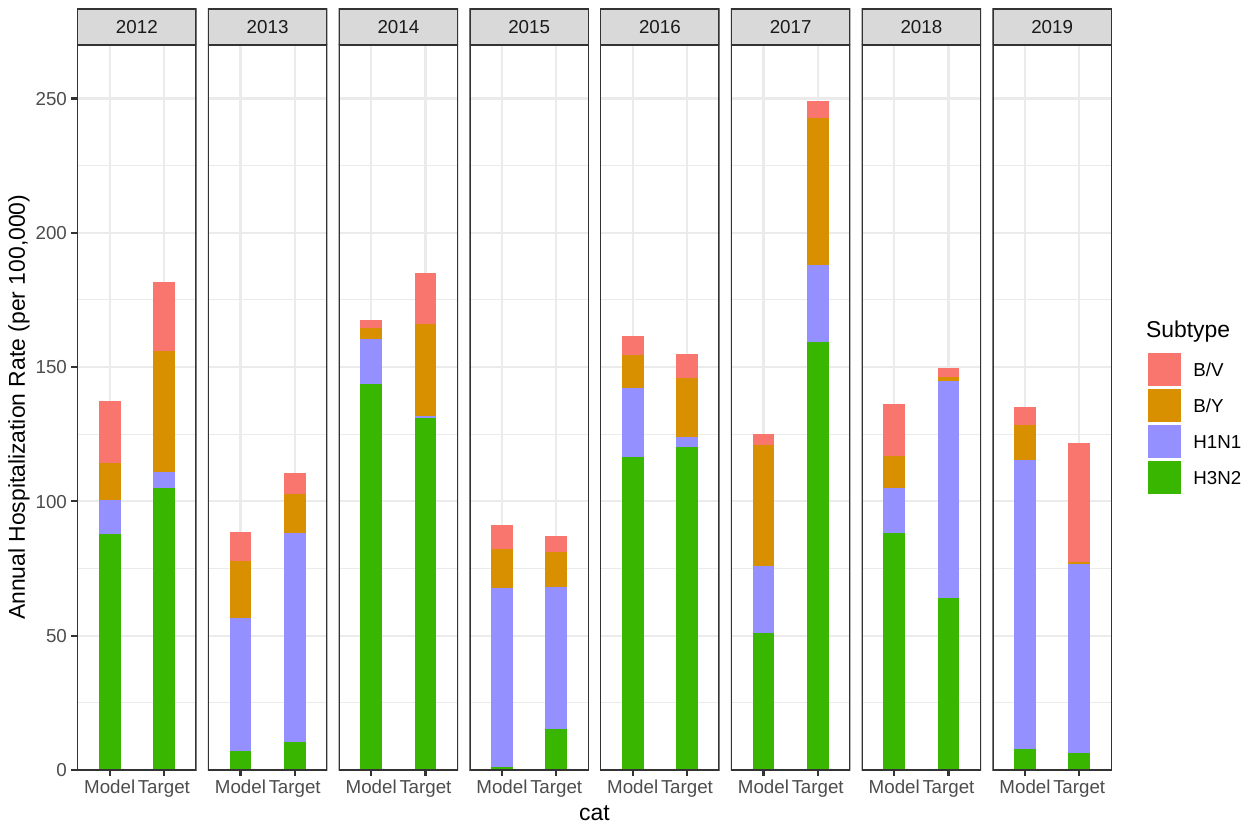

**Fig S3. Comparison of simulated and observed influenza strain composition during 2012/13 – 2019/20 seasons.**
